# Feasibility of Endothelial Cell Isolation from Routine Coronary Function Testing in ANOCA Patients

**DOI:** 10.64898/2026.04.09.26350551

**Authors:** Elize A.M. de Jong, Daniek M.C. Kapteijn, Mark A. Daniels, Timo Nijkamp, Peter D. Zalewski, John F. Beltrame, Peter Damman, Mete Civelek, Ernest Diez Benavente, Tim P. van de Hoef, Hester M. den Ruijter

**Affiliations:** Laboratory of Experimental Cardiology, University Medical Centre Utrecht, Utrecht University, Utrecht, The Netherlands; Department of Cardiology, Division Heart and Lungs, University Medical Centre Utrecht, Utrecht University, Utrecht, The Netherlands; Basil Hetzel Institute for Translational Health Research, Adelaide Medical School, College of Health, Adelaide University; Basil Hetzel Institute for Translational Health Research, Woodville, South Australia, Australia; Adelaide Medical School, Adelaide University, Adelaide, South Australia, Australia; Central Adelaide Local Health Network, Department of Cardiology, Adelaide, South Australia, Australia; Department of Cardiology, RadboudUMC, Radboud University, Nijmegen, The Netherlands; Department of Anesthesiology and Perioperative Medicine, University of California, Los Angeles, California, USA

**Keywords:** Vascular dysfunction, Coronary microvascular dysfunction, Primary cell culture, Translational research

## Abstract

**Background:** Angina with nonobstructive coronary arteries (ANOCA) is a heterogeneous condition encompassing distinct endotypes representing different underlying pathophysiological mechanisms. Endothelial dysfunction is considered a central hallmark of ANOCA. However, studying patient-derived endothelial cells (ECs) remains challenging due to the limited availability of disease-specific endothelial samples. We therefore aimed to assess the feasibility of isolating and culturing ECs from catheterization material obtained during routine coronary function testing in ANOCA patients.

**Methods:** Catheterization material was collected from 79 ANOCA patients (84% female, age 58±10 years) undergoing coronary function testing. ECs were isolated, expanded and characterized using immunostaining, flow cytometry, gene expression profiling and functional assays.

**Results:** EC isolation was successful in 43% of cases and resulted in 34 primary EC cultures that were expanded up to passage 10. Isolation success was independent of clinical or procedural characteristics. Isolated cells exhibited typical EC morphology and expressed EC markers confirmed by immunostaining, flow cytometry and gene expression analyses. EC marker gene expression remained largely stable over passages. However, stress- and defense-related gene expression programs increased over time, while proliferation-related processes decreased. Functional assays demonstrated that the coronary catheterization-derived ECs showed typical properties of wound healing, angiogenesis, activation responses upon stimuli and monocyte adhesion.

**Conclusions:** This study demonstrates the feasibility of isolating and expanding ECs directly from catheterization material collected during routine coronary function testing in ANOCA patients. These patient-derived ECs retain characteristic endothelial features and functionality. This approach offers primary EC cultures to study the mechanisms underlying endothelial dysfunction in ANOCA.

**Graphic Abstract:** 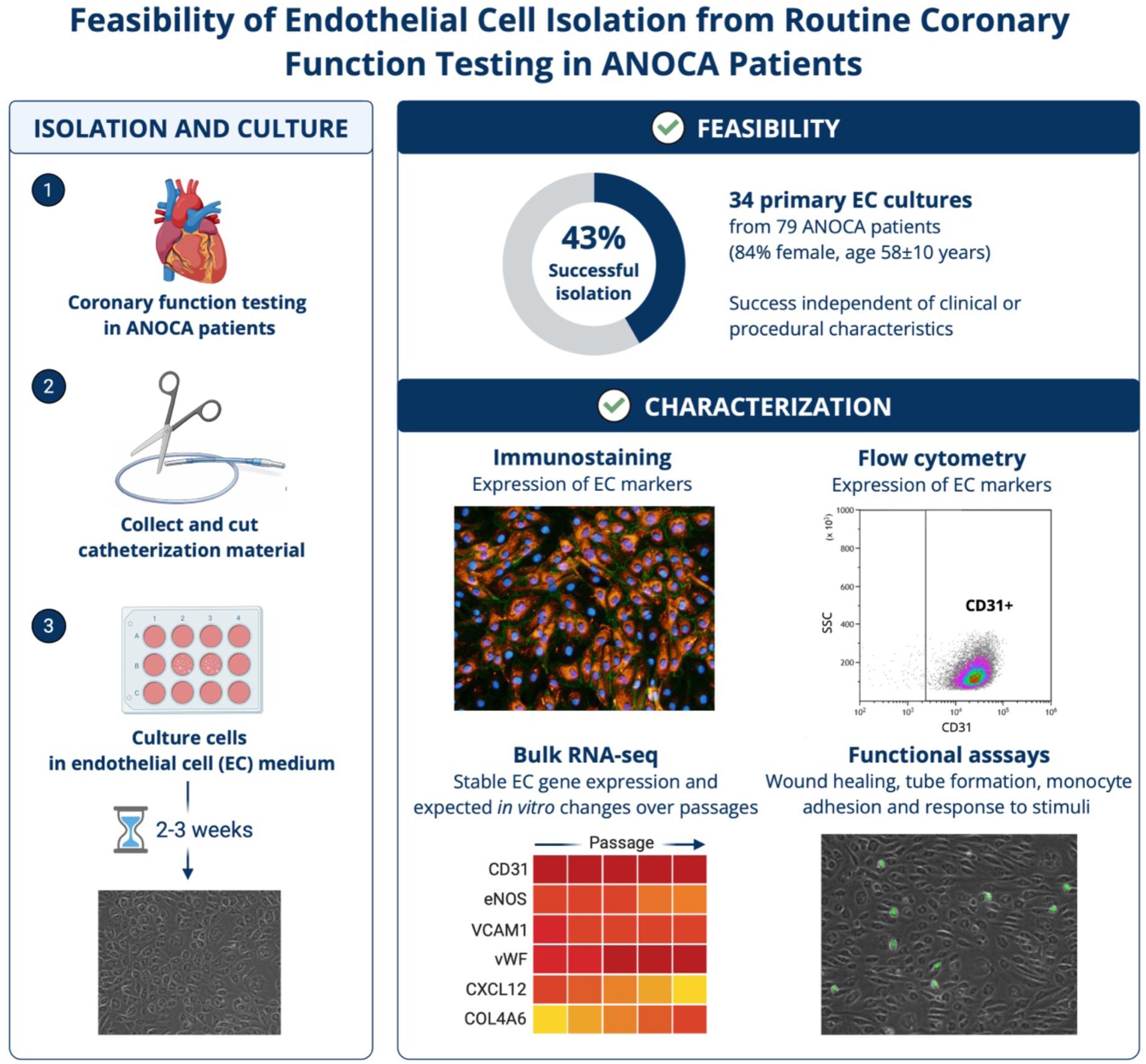

**Perspectives:** *Clinical perspective:* What is new?

- We established a method to isolate and culture endothelial cells from routine coronary catheterization material in patients with ANOCA, enabling direct study of patient-specific endothelial dysfunction.
- The patient-derived endothelial cells exhibit characteristic morphology, express canonical endothelial markers and retain functional properties consistent with endothelial physiology. What are the clinical implications?

- This approach provides a clinically relevant platform to investigate mechanisms underlying ANOCA and may support the development of personalized diagnostic and therapeutic strategies.
- Broader application of this method could facilitate translational research in other vascular pathologies where access to endothelial tissue is limited.

## Introduction

Angina with nonobstructive coronary arteries (ANOCA) is a highly prevalent yet clinically challenging condition among patients undergoing coronary angiography. The syndrome encompasses coronary vasomotor dysfunction with distinct endotypes that reflect diverse underlying pathophysiological mechanisms. These endotypes include coronary vasospasm, resulting from increased vasoconstriction in either the epicardial arteries or the microvasculature, and coronary microvascular dysfunction, characterized by impaired vasodilatory capacity of the coronary microcirculation^1–3^. Effective, evidence-based therapies are currently limited, as the precise mechanisms driving these endotypes remain incompletely understood. Nonetheless, despite the limited mechanistic knowledge, there is broad consensus that endothelial dysfunction represents a central hallmark of coronary vasomotor dysfunction in ANOCA^4,5^.

Endothelial dysfunction is characterized by an imbalance between vasoconstrictive and vasodilatory signaling, resulting in impaired endothelium-dependent vasodilation. Beyond vascular tone dysregulation, it also involves disturbances in key homeostatic processes, including inflammation, vascular remodeling, thrombosis and barrier integrity^6,7^. These functional alterations reflect dysregulation of molecular programs within the vascular wall. In this context, our recent work using sex-stratified gene regulatory network analyses in human cardiovascular disease has demonstrated that vascular transcriptional programs are fundamentally sex-specific, identifying female-specific networks and key driver genes^8,9^. These findings suggest that endothelial and vascular cell phenotypes are shaped by sex-dependent regulatory mechanisms, which may influence susceptibility to coronary vasomotor dysfunction. Given that ANOCA and vasomotor dysfunction disproportionately affect women, incorporating a sex- and patient-specific perspective into studies of endothelial dysfunction is likely essential to fully elucidate its underlying mechanisms.

Experimental research on endothelial dysfunction commonly relies on established *in vitro* models, such as human endothelial cell (EC) lines, primary ECs and advanced three-dimensional or microfluidic culture systems that recapitulate aspects of vascular physiology^10^. While these systems have greatly enhanced our understanding of endothelial biology, they cannot fully capture the complexity and heterogeneity of patient-specific vascular pathology. Consequently, the limited availability of disease-specific endothelial samples has constrained efforts to study patient-derived ECs and their mechanistic contribution to vasomotor dysfunction development.

Previously, we described a method to harvest human arterial ECs from catheterization material obtained during clinically indicated diagnostic coronary angiography^11^. While this approach demonstrated the feasibility of isolating patient-derived ECs, its application remained limited to short-term characterization, without subsequent propagation in culture. This is an important goal as prior work from our group has shown that primary vascular-derived cells can retain disease-specific molecular and functional characteristics in culture^12,13^, supporting their use as a relevant model to experimentally investigate gene candidates and disease mechanisms. Therefore, we refined the established approach and present a method to isolate and culture ECs from catheterization material obtained during routine diagnostic procedures. This method establishes a platform to investigate disease-specific endothelial mechanisms underlying ANOCA in a controlled experimental setting.

## Methods

### Study design and participants

ECs were isolated and cultured from catheterization material (guiding catheter, guide wire, PressureWire X (Abbott, USA) and RayFlow microcatheter (Hexacath, France)) obtained directly after diagnostic coronary function testing in patients with ANOCA. Samples were collected in the University Medical Center Utrecht as part of the ENDOTYPE biobank (TCbio 24-203), for which written informed consent was obtained from all participants prior to the procedure. One eligibility criterion for inclusion in the ENDOTYPE biobank was participation in the NL-CFT registry (21-473)^14^, enabling linkage of clinical and procedural data to corresponding biological samples. Ethical approval for the biobank was granted by the Biobank Committee under the responsibility of the Medical Ethical Committee of the University Medical Center Utrecht. All procedures were conducted in accordance with the Declaration of Helsinki. Data supporting the findings of this study are available from the corresponding author upon reasonable request.

### Isolation and culture of endothelial cells from catheterization material

ECs were isolated from catheterization material obtained immediately after the coronary function test. Catheters and wires were cut into short segments of approximately 5 centimeters and placed in 4°C complete EC medium (Endothelial Cell Basal MV, phenol-red free (PromoCell, REF#C-22225) supplemented with EC Growth Medium MV SupplementPack (PromoCell, REF#C-39220) and 1% penicillin/streptomycin (Fisher Scientific, REF#15-140-122)). Next, catheter wires were transported on ice to the laboratory. To release adherent ECs from the catheter wires, they were rotated in complete EC medium for 10 min at 4 °C, after which the catheter wires were removed. Cells were pelleted by centrifugation (330 × g, 5 min, room temperature (RT)), resuspended in complete EC medium, and plated on 0.1% gelatin–coated 12-well plates. Cultures were maintained at 37 °C, 5% CO₂, and refreshed three times weekly. Red blood cells and non-adherent cells were progressively removed by medium changes, and endothelial colonies became visible within 1–3 weeks depending on donor material.

At ∼80% confluence, cells were detached using Accutase (Innovative Cell Technologies, REF#AT104) and passaged sequentially into 6-well plates and T75 flasks pre-coated with 0.1% gelatin. Cells were cultured up to passage 10 (P10). At each passage, aliquots were used for downstream applications including flow cytometry, RNA isolation, or cryopreservation. For freezing, cells were resuspended in freezing medium (80% complete EC medium, 20% DMSO), cooled at –80 °C in a controlled-rate freezing container, and transferred to liquid nitrogen storage.

### Immunofluorescence

Endothelial cells derived from coronary catheterization material (coronary catheterization-derived ECs; CC-ECs) were cultured on 0.1% gelatin-precoated glass slides. EC medium was aspirated and ECs were fixed with 4% paraformaldehyde (Santa Cruz Technologies; REF#sc-281692) for 15 min at RT, followed by washing with 1x phosphate-buffered saline (PBS; Gibco; REF#10010056). Permeabilization was performed using 0.1% TritonX-100 in 1x PBS for 10 minutes, after which the cells were washed and blocked with 10% normal goat serum (NGS, REF#50197Z) in 1x PBS for 60 min, RT. Subsequently, the cells were incubated overnight at 4°C with the following primary antibodies diluted in 1% bovine serum albumin in PBS (PBSA): polyclonal goat anti-human CD31 (0.67µg/mL; Santa Cruz Technologies; REF#sc-376764), monoclonal mouse anti-human VE-Cadherin (0.5 µg/mL; Santa Cruz Technologies; REF#SC-9989), polyclonal rabbit anti-human von Willebrand Factor (vWF, 14 µg/mL; R&D systems; REF#A0082) The next day, slides were washed three times with 1xPBS and incubated with the corresponding fluorophore-conjugated secondary antibodies: goat anti-rabbit Alexa Fluor 555 (2.5 µg/mL; Fisher Scientific; REF#A21428) and goat anti-mouse Alexa Fluor 488 (2.5 µg/mL Fisher Scientific; REF#A11001). After three additional PBS washes, nuclei were counterstained with Hoechst (1:10.000 in PBS) for 3min at RT. Slides were then washed once more with PBS and mounted with Fluoromount-G (SouthernBiotech; 0100-01). Slides were stored at 4°C in the dark for short-term use or at - 20°C for long-term preservation. Imaging was performed using an Olympus BX53 fluorescence widefield microscope.

### Flow cytometry characterization of CC-ECs

CC-ECs were harvested at different passages and resuspended in 50 μl Fluorescence-Activated Cell Sorting (FACS) buffer (94.8% PBS (Gibco, REF#10010056), supplemented with 5% fetal bovine serum (FBS; Corning, REF#35-079-CV) and 0.2% EDTA (Sigma-Aldrich, REF#E4884-500G)). Cells were stained with fluorescently labeled antibodies against CD144 (VE-cadherin), CD31 (PECAM1), CD34, CD14, CD45 and CD41. Detailed information is provided in **Supplemental Table S1**. Antibodies were prepared at the indicated working concentrations in PBS to a final volume of 100 μl and incubated for 30 minutes at 4°C. Cells were then washed with 2 ml FACS buffer and stained with Zombie NIR viability dye diluted 1:1000 in PBS, to exclude non-viable cells (**Supplemental Table S1**). After incubation and washing, ECs were resuspended in 250 µl FACS buffer and measured on a CytoFLEX flow cytometer (Beckman Coulter). Data was analyzed using Kaluza Analysis Software (version 2.1; Beckman Coulter). Reference populations included human dermal microvascular ECs (HDMVEC; iXCells Biotechnologies, REF#10HU-019), human cardiac microvascular ECs (HCMEC; Sigma-Aldrich, REF#C-12285), human coronary artery ECs (HCAEC; Lonza, REF#CC-2585), human plaque myofibroblasts^12^ and mesenchymal stem cells (MSC; Cell Therapy Facility, University Medical Center Utrecht; code: MSC053P3_AL-MSC071P3_R).

### RNA isolation

Total RNA was isolated using TriPure Isolation Reagent (Sigma-Aldrich, REF#11667165001). Cells were lysed in 200 µL of TriPure and incubated for 5 min at RT. Chloroform (Sigma-Aldrich, REF#32211-1L) was added, samples were mixed vigorously and incubated for 2–3 min at RT. Next, samples were centrifuged at 12000 × g for 15 min at 4 °C. Following phase separation, the upper aqueous phase was carefully transferred to a fresh RNase-free tube. Thereafter, RNA was precipitated by adding isopropanol (Sigma-Aldrich, REF#33539-2.5L-R) and visualized by adding 0.5 µL RNase-free GlycoBlue Coprecipitant (ThermoScientific, REF#AM9516). Samples were mixed by inversion and incubated overnight at −20 °C to maximize RNA-recovery. The next day, precipitated RNA was pelleted by centrifugation at 12000 × g for 10 min at 4 °C. The pellet was washed twice with 75% ethanol (EMD Millipore, REF#1.00983.2500). Each wash was followed by centrifugation at 7500 × g for 5 minutes at 4 °C. After removal of residual ethanol, pellets were air-dried for 10-30 minutes and dissolved in RNase-free water. RNA concentration was quantified using the Qubit Fluorometer (Thermo Fisher Scientific, REF#Q32855) following the manufacturer’s instructions.

### Bulk RNA sequencing

Bulk 3’ RNA sequencing was performed by Single Cell Discoveries (Utrecht, The Netherlands) according to their standard bulk RNA-sequencing workflow. Purified RNA was reverse transcribed using sample-specific barcodes to generate cDNA, pooled and amplified via in vitro transcription. Quality control of amplified RNA and final cDNA libraries was performed using TapeStation and Qubit. Libraries were sequenced on an Illumina platform with a target depth of approximately 10 million reads per sample. Following sequencing, demultiplexed FASTQ files were mapped to the human reference genome (GRCh38) using STARsolo (v2.7.11b), and gene-level read counts were generated for downstream analysis. Length-based normalization was not required due to 3’-end capture.

### Functional characterization of CC-ECs

Functional characterization of CC-ECs was performed using complementary *in vitro* assays to assess cell migration, angiogenesis, inflammatory activation and monocyte adhesion. Primary human umbilical vein ECs (HUVECs) were used throughout as a reference EC type to enable comparison with established EC functional responses.

#### Wound healing assay: cell migration

For cell migration analysis, confluent EC monolayers were seeded in 24-well plates. For each donor, ECs were plated in three independent wells, and the scratch assay was performed in triplicate. Upon reaching 100% confluency, the wells were scratched with a sterile 200 µl pipette tip to generate a linear wound. Detached cells were removed by washing with PBS, and cultures were maintained in complete EC medium. Images of the wound area were acquired every hour using the Omni live cell imager (CytoSMART). Wound closure was quantified as the reduction in scratch area over time, using ImageJ software.

#### Angiogenesis assay: tube formation

Angiogenic capacity of CC-ECs was assessed by performing a tube formation assay using growth factor-reduced Matrigel (Corning, REF#354230). Ibidi plates were coated with 10µl Matrigel per well and incubated for 30 minutes at 37 °C to allow polymerization. ECs were resuspended in complete EC medium and seeded at a density of 5 × 10^3^ cells per well. Cells were incubated at 37°C, 5% CO₂ for 24 hours, after which tube-like network formation was visualized using an inverted phase-contrast microscope. The assay was performed with four replicates per donor, and the mean of these replicates was used for quantitative analysis of tube length, number of nodes, and meshes was performed using ImageJ (Angiogenesis Analyzer plugin^15^).

#### Flow cytometry characterization of activated ECs

To evaluate whether CC-ECs respond to inflammatory stimuli similarly to established EC models, ECs were activated using different stimuli. 50.000 ECs were seeded per well in a 24 well plate, to get 80% confluency. The next day, complete EC medium was changed to starvation medium with either 10ng/ml transforming growth factor-beta (TGF-β, Peprotech, REF#100-35-10ng), 10ng/ml tumor necrosis factor-alpha (TNFα, Miltenyi Biotec, REF#130-094-018) and 5ng/ml interleukin-1 beta (IL-1β, R&D Systems, REF#201-LB-005/CF) and incubated for 24 hours at 37°C, 5%CO_2_. Afterwards, flow cytometric analysis of the stimulated CC-ECs were performed as described above using fluorescently labeled antibodies to assess EC-specific markers **(Supplemental Table S1)**.

#### Monocyte adhesion assay

To investigate interactions between (activated) ECs and monocytes (THP1 cells), we performed a fluorescence-based adhesion assay, using the ibidi flow system. ECs were cultured to approximately 80% confluency prior to flow experiments. A total of 3 × 10^5^ ECs were seeded into µ-Slide I Luer slides (0.4 mm channel height; ibidi, REF#80176) and allowed to attach for 2 hours at 37 °C, 5% CO₂ before connection to the ibidi pump system. Each µ-slide was connected to an ibidi perfusion set (yellow/green, 50 cm, ID 1.6 mm; REF#10964), pre-filled with 12 ml complete EC medium. ECs were cultured overnight under physiological flow conditions (5.8 mbar pressure, shear stress: 4.0 dyn/cm², flow rate: 3.04 ml/min, shear rate: 400 s⁻¹, unidirectional: 20 s, oscillating: 0.5 s). The next day, complete EC medium was replaced with 12 ml starvation medium (Endothelial Cell Basal MV, phenol-red free (PromoCell, REF#C-22225) supplemented with 5% charcoal-stripped FBS (Thermofischer Scientific, REF#12676029)). ECs were either stimulated with 10 ng/ml TNFα for 5 hours at 37 °C, 5% CO₂ or left untreated (control). In parallel, THP-1 monocytes were harvested and labeled using the CellTrace CFSE Cell proliferation kit (ThermoFischer Scientific, REF#C34554). Subsequently, 1 × 10^6^ fluorescently labeled monocytes were introduced into each perfusion system and circulated for 2 hours under the same flow conditions as described above. Thereafter, µ-slides were disconnected and gently rinsed with 1x PBS to remove non-adherent monocytes. Morphological images were made using the Olympus BX53 fluorescence widefield microscope and analyzed using ImageJ software.

### Statistical analysis

Normally distributed continuous variables, as assessed by the Shapiro-Wilk test, are presented as mean ± SD, while non-normally distributed data are shown as median (IQR). Categorical variables are reported as number (%). For the functional assays, differences in wound closure rates (defined by linear regression slopes), tube formation parameters, expression of EC markers and flow-based monocyte adhesion were compared between CC-ECs and HUVECs using the Wilcoxon rank-sum test.

Bulk RNA-seq counts were imported and preprocessed in R using DESeq2. Genes with <10 counts in at least 3 samples were removed. Variance-stabilizing transformation was applied for principal component analysis and EC-defining genes over passages, while raw counts were used for differential expression analysis with passage number modelled as continuous variable. Genes with an adjusted p-value <0.05 and absolute log2 fold change greater than 0.25 were considered significant. To generate the volcano plot, the EnhancedVolcano package was used to generate the volcano plot. Gene set enrichment analysis was performed on ranked log2 fold changes using clusterProfiler’s gseGO() function, with pathways classified as up- or downregulated based on the normalized enrichment score (NES). In addition, bulk expression profiles were projected onto a reference single-cell RNA-seq dataset^16^ using SingleR, which was run with default settings on log-normalized counts from the reference; similarity scores were visualized as heatmaps.

## Results

### Patient population

Catheterization material was collected from 79 ANOCA patients undergoing coronary function testing (**Figure 1A**). Baseline characteristics are shown in **Table 1**. The mean age was 58 ± 10 years and 84% of patients were female. Arterial access was predominantly obtained via the radial artery (86%). When radial access was not feasible, femoral (13%) or ulnar (1%) access was used. Acetylcholine spasm provocation testing resulted in a diagnosis of epicardial spasm in 20 patients (25%), microvascular spasm in 22 patients (28%) and combined epicardial and microvascular spasm in 6 patients (8%). An inconclusive spasm test (not meeting the COVADIS criteria) was observed in 16 patients (20%), while 15 patients (19%) had a negative spasm test. Overall, patients had a mean fractional flow reserve of 0.95 ± 0.03, a median coronary flow reserve of 3.0 (2.6 – 3.6) and a median microvascular resistance reserve of 3.4 (2.8 – 4.0). Based on a coronary flow reserve <2.5, coronary microvascular dysfunction was diagnosed in 17 patients (22%). Cardiovascular medication use was common in this cohort.

**Figure 1.**
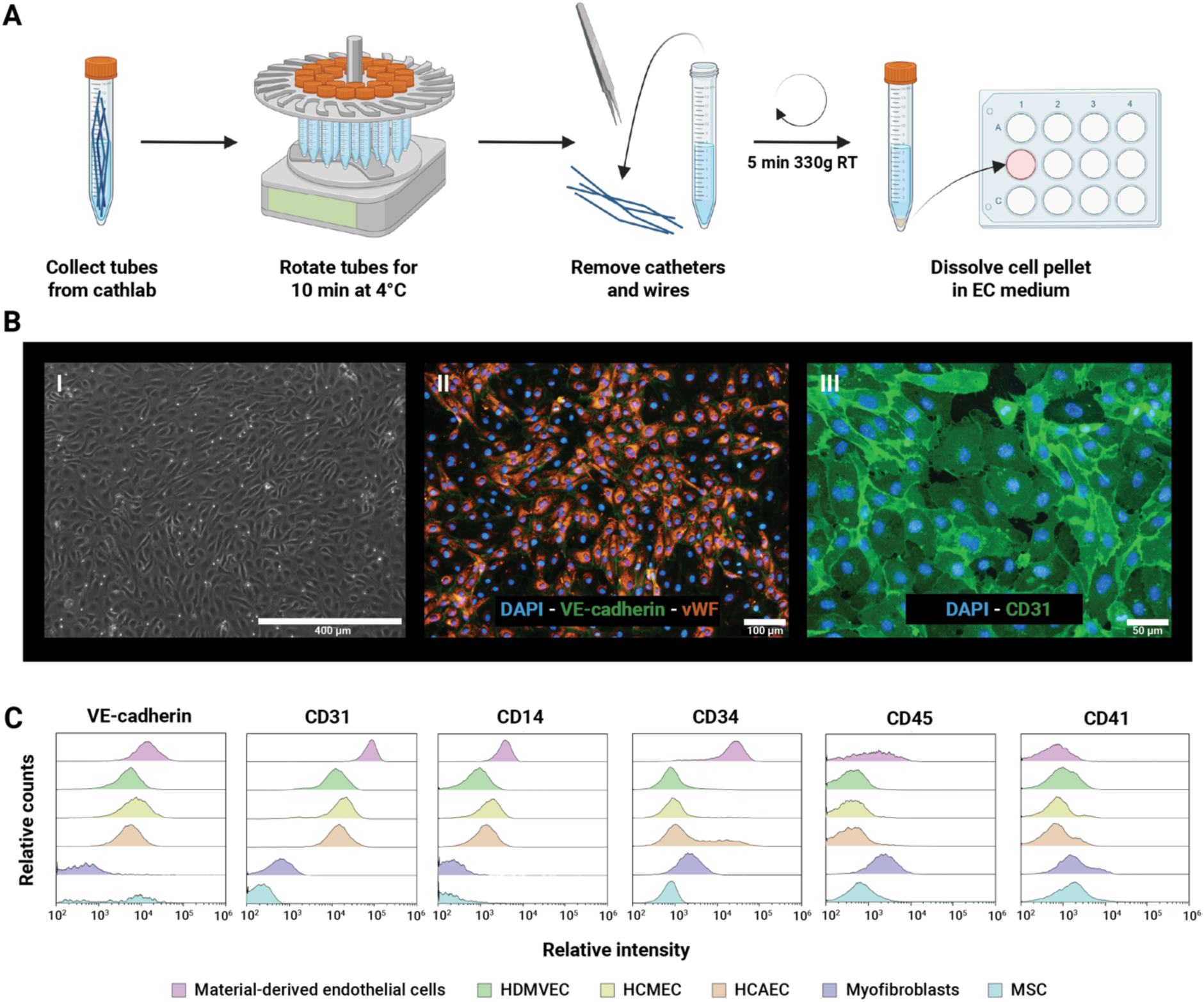
Characterization of coronary catheterization-derived endothelial cells (CC-ECs). A) Method to isolate and culture ECs from catheterization material used during coronary function testing. B) Representative morphology (I, passage 0) and immunofluorescence images of cultured ECs (II and III, passage 5) showing positivity for VE-cadherin (II), von Willebrand Factor (vWF) (II) and CD31 (III). C) Flow-cytometric characterization of cultured ECs (passage 1) in comparison with multiple reference cell populations, including human dermal microvascular ECs (HDMVEC), human cardiac microvascular ECs (HCMEC), human coronary artery ECs (HCAEC), human plaque myofibroblasts and mesenchymal stem cells (MSC). The plotted histograms depict the ‘relative counts’ on the y-axis and the ‘relative intensity’ on the x-axis

**Table 1.** Baseline characteristics of patients from whom endothelial cells were isolated.

|  | Total cohort<br>(N = 79) | Successful<br>(n = 34) | Unsuccessful<br>(n = 45) |
| --- | --- | --- | --- |
| **Age, years** | 58 ± 10 | 57 ± 8 | 58 ± 11 |
| **Female sex** | 66 (84%) | 30 (88%) | 36 (80%) |
| **BMI, kg/m<sup>2</sup>** | 27.6 ± 4.7 | 27.0 ± 4.9 | 28.1 ± 4.6 |
| **Smoking** |  |  |  |
| Never smoked | 33 (42%) | 12 (35%) | 21 (47%) |
| Current smoker | 6 (8%) | 3 (9%) | 3 (7%) |
| Former smoker | 22 (28%) | 8 (24%) | 14 (31%) |
| Unknown | 18 (23%) | 11 (32%) | 7 (16%) |
| **Diabetes Mellitus** |  |  |  |
| Yes | 7 (9%) | 2 (6%) | 5 (11%) |
| No | 69 (87%) | 30 (88%) | 39 (87%) |
| Unknown | 3 (4%) | 2 (6%) | 1 (2%) |
| **Hypertension** |  |  |  |
| Yes | 35 (44%) | 13 (38%) | 22 (49%) |
| No | 43 (54%) | 20 (59%) | 23 (51%) |
| Unknown | 1 (1%) | 1 (3%) | 0 (0%) |
| **Hypercholesterolemia** |  |  |  |
| Yes | 29 (37%) | 10 (29%) | 19 (42%) |
| No | 41 (52%) | 18 (53%) | 23 (51%) |
| Unknown | 9 (11%) | 6 (18%) | 3 (7%) |
| **Arterial access** |  |  |  |
| Radial artery | 68 (86%) | 31 (91%) | 37 (82%) |
| Femoral artery | 10 (13%) | 2 (6%) | 8 (18%) |
| Ulnar artery | 1 (1%) | 1 (3%) | 0 (0%) |
| **Acetylcholine spasm provocation test** |  |  |  |
| No spasm | 15 (19%) | 8 (24%) | 7 (16%) |
| Inconclusive test | 16 (20%) | 5 (15%) | 11 (24%) |
| Epicardial spasm | 20 (25%) | 8 (24%) | 12 (27%) |
| Microvascular spasm | 22 (28%) | 10 (29%) | 12 (27%) |
| Combined epicardial and microvascular spasm | 6 (8%) | 3 (9%) | 3 (7%) |
| **Absolute flow measurements** |  |  |  |
| FFR | 0.95 ± 0.03 | 0.95 ± 0.02 | 0.95 ± 0.04 |
| CFR | 3.0 (2.6 – 3.6) | 3.1 (2.8 – 4.0) | 2.9 (2.5 – 3.4) |
| MRR | 3.4 (2.8 – 4.0) | 3.4 (3.0 – 4.2) | 3.3 (2.7 – 4.0) |
| **Diagnosis of CMD based on CFR <2.5** | 17 (22%) | 6 (18%) | 11 (24%) |
| **Cardiovascular medication use** |  |  |  |
| Antiplatelet therapy | 37 (46%) | 15 (44%) | 22 (49%) |
| Lipid-lowering drugs | 41 (52%) | 21 (62%) | 20 (44%) |
| Calcium-channel blockers | 51 (65%) | 21 (62%) | 30 (67%) |
| Nitrates | 47 (59%) | 21 (62%) | 26 (58%) |
| Beta-blockers | 20 (25%) | 9 (26%) | 11 (24%) |
| ACE-inhibitors or ARBs | 23 (29%) | 8 (24%) | 15 (33%) |

Values are presented as mean ± SD, median (IQR) or n (%). Abbreviations: ACE, angiotensin-converting enzyme; ARB, angiotensin receptor blocker; BMI, body mass index; CFR, coronary flow reserve; CMD, coronary microvascular dysfunction; FFR, fractional flow reserve; MRR, microvascular resistance reserve.

|  | **Total cohort**<br>**(N = 79)** | **Successful**<br>**(n = 34)** | **Unsuccessful**<br>**(n = 45)** |
| --- | --- | --- | --- |
| **Age, years** | 58 ± 10 | 57 ± 8 | 58 ± 11 |
| **Female sex** | 66 (84%) | 30 (88%) | 36 (80%) |
| **BMI, kg/m<sup>2</sup>** | 27.6 ± 4.7 | 27.0 ± 4.9 | 28.1 ± 4.6 |
| **Smoking** |  |  |  |
| Never smoked | 33 (42%) | 12 (35%) | 21 (47%) |
| Current smoker | 6 (8%) | 3 (9%) | 3 (7%) |
| Former smoker | 22 (28%) | 8 (24%) | 14 (31%) |
| Unknown | 18 (23%) | 11 (32%) | 7 (16%) |
| **Diabetes Mellitus** |  |  |  |
| Yes | 7 (9%) | 2 (6%) | 5 (11%) |
| No | 69 (87%) | 30 (88%) | 39 (87%) |
| Unknown | 3 (4%) | 2 (6%) | 1 (2%) |
| **Hypertension** |  |  |  |
| Yes | 35 (44%) | 13 (38%) | 22 (49%) |
| No | 43 (54%) | 20 (59%) | 23 (51%) |
| Unknown | 1 (1%) | 1 (3%) | 0 (0%) |
| **Hypercholesterolemia** |  |  |  |
| Yes | 29 (37%) | 10 (29%) | 19 (42%) |
| No | 41 (52%) | 18 (53%) | 23 (51%) |
| Unknown | 9 (11%) | 6 (18%) | 3 (7%) |
| **Arterial access** |  |  |  |
| Radial artery | 68 (86%) | 31 (91%) | 37 (82%) |
| Femoral artery | 10 (13%) | 2 (6%) | 8 (18%) |
| Ulnar artery | 1 (1%) | 1 (3%) | 0 (0%) |
| **Acetylcholine spasm provocation test** |  |  |  |
| No spasm | 15 (19%) | 8 (24%) | 7 (16%) |
| Inconclusive test | 16 (20%) | 5 (15%) | 11 (24%) |
| Epicardial spasm | 20 (25%) | 8 (24%) | 12 (27%) |
| Microvascular spasm | 22 (28%) | 10 (29%) | 12 (27%) |
| Combined epicardial and microvascular spasm | 6 (8%) | 3 (9%) | 3 (7%) |
| **Absolute flow measurements** |  |  |  |
| FFR | 0.95 ± 0.03 | 0.95 ± 0.02 | 0.95 ± 0.04 |
| CFR | 3.0 (2.6 – 3.6) | 3.1 (2.8 – 4.0) | 2.9 (2.5 – 3.4) |
| MRR | 3.4 (2.8 – 4.0) | 3.4 (3.0 – 4.2) | 3.3 (2.7 – 4.0) |
| **Diagnosis of CMD based on CFR <2.5** | 17 (22%) | 6 (18%) | 11 (24%) |
| **Cardiovascular medication use** |  |  |  |
| Antiplatelet therapy | 37 (46%) | 15 (44%) | 22 (49%) |
| Lipid-lowering drugs | 41 (52%) | 21 (62%) | 20 (44%) |
| Calcium-channel blockers | 51 (65%) | 21 (62%) | 30 (67%) |
| Nitrates | 47 (59%) | 21 (62%) | 26 (58%) |
| Beta-blockers | 20 (25%) | 9 (26%) | 11 (24%) |
| ACE-inhibitors or ARBs | 23 (29%) | 8 (24%) | 15 (33%) |

Values are presented as mean ± SD, median (IQR) or n (%). Abbreviations: ACE, angiotensin-converting enzyme; ARB, angiotensin receptor blocker; BMI, body mass index; CFR, coronary flow reserve; CMD, coronary microvascular dysfunction; FFR, fractional flow reserve; MRR, microvascular resistance reserve.

### Establishment of EC cultures from catheterization material

Among the 79 ANOCA patients who underwent coronary function testing, EC cultures were successfully established in 34 cases (43%) (**Table 1**). In these successful isolations, the first endothelial colonies appeared after approximately two to three weeks in primary 12-well cultures. Once colonies reached confluence, cultures were expanded into 6-well plates after four to five weeks (passage 1) and subsequently cultured up to passage 10. In the remaining 45 cases (57%), no EC colonies were observed within the first two to three weeks, after which the cultures were discontinued. Baseline clinical and procedural characteristics were comparable between patients with successful and unsuccessful isolations (**Table 1**). We tested whether any of the clinical or procedural characteristics were predictors of isolation success and identified through univariate logistic regression analysis that none of the tested variables associated with EC outgrowth success (**Supplemental Table S2**). To understand which type of catheterization material was most effective as a resource, we tested different types in a subset of 59 samples. We tested the guiding catheter, guide wire, PressureWire X and RayFlow microcatheter. Successful EC cultures were consistently obtained from devices entering the coronary artery (guide wire, PressureWire X or Rayflow microcatheter), but never from the guiding catheter, suggesting a likely coronary origin in most cases.

For characterization of the isolated ECs, we used cells derived from different passages and donors, as summarized in **Supplemental Table S3**.

### Characterization of CC-ECs

CC-ECs displayed a characteristic endothelial morphology two to three weeks after isolation and throughout early culture as reflected by a homogeneous monolayer of cells with the typical cobblestone-like appearance (**Figure 1B, I**). The proliferative capacity of the resulting EC cultures was comparable to that of HUVECs^17^. To further verify endothelial identity, we stained the cells with immunofluorescence for canonical EC markers. CC-ECs showed positivity for VE-cadherin, vWF and CD31, confirming the presence of key junctional and cytoplasmic proteins characteristic of functional endothelium (**Figure 1B, II-III**). Flow cytometric analysis at passage 1 supported the EC origin (**Figure 1C**). Our CC-ECs expressed high levels of VE-cadherin and CD31, consistent with endothelial reference populations, including HDMVECs, HCMECs and HCAECs. CD14 expression was detected in all ECs, with variable levels across different endothelial subsets, which is thought to reflect context-dependent activation states^18^. Moreover, CD34 expression was higher in CC-ECs (passage 1) compared to reference EC populations (higher passages). However, CD34 decreased with continued passaging in our ECs (**Supplemental Figure S1**), consistent with previous studies showing that CD34 is expressed in freshly isolated ECs but diminishes upon serial passaging *in vitro*^19,20^. Furthermore, as expected for endothelial lineage cells, the populations were negative for the hematopoietic marker CD45 and the platelet lineage marker CD41, indicating the absence of contamination by immune or megakaryocytic cells (**Figure 1C**).

### Transcriptional characterization and stability of CC-ECs in culture

To evaluate changes in gene expression in CC-ECs in culture over time, we performed bulk RNA sequencing across multiple passages in the first nine successfully established CC-EC cultures. These cultures were not selected based on cellular phenotype or donor clinical characteristics. Principal component analysis showed donor-specific clusters irrespective of passage number, indicating a stable transcriptional profile within each donor (**Figure 2A**). To confirm endothelial identity at the transcriptomic level, we projected the bulk RNA profiles onto an established single-cell RNA sequencing reference atlas from coronary arteries^16^. Here, the CC-EC transcriptomes localized specifically to the EC cluster, as shown by the UMAP embedding and corresponding cell-type annotation (**Figure 2B**). Expression levels of genes involved in key endothelial functions, including vascular barrier integrity (*CD31, VE-cadherin*), vascular tone regulation (*eNOS, EDN1*), inflammatory responses (*ICAM1, VCAM1*) and coagulation (*vWF, PROCR*), remained stable across passages (**Figure 2C**), suggesting functional preservation of these endothelial programs during *in vitro* culture. Finally, we modelled passage number as a continuous variable and identified 675 genes that significantly decreased (Log2FC <0.25; e.g., *PCLAF, CXCL12, TMEM100, RFC3, SULF1, NRK*) and 472 genes that significantly increased (Log2FC >0.25; e.g., *CD82, COL4A6, ICAM2, PSMB8, CCND1, CXCL11*) with prolonged culture (adjusted *p*<0.05) (**Figure 2D; Supplemental Table S4**). Pathway enrichment analysis of passage-associated genes revealed downregulation of cell division and proliferation-related pathways, suggesting that transcriptomic changes with passaging primarily reflect shifts in proliferative state rather than loss of endothelial identity. Conversely, antiviral and defense-related pathways, including negative regulation of viral genome replication, response to virus and lysosomal/endopeptidase activity, were higher upon longer culture, indicating increased stress- and defense-related processes over passages (**Figure 2E**).

**Figure 2.**
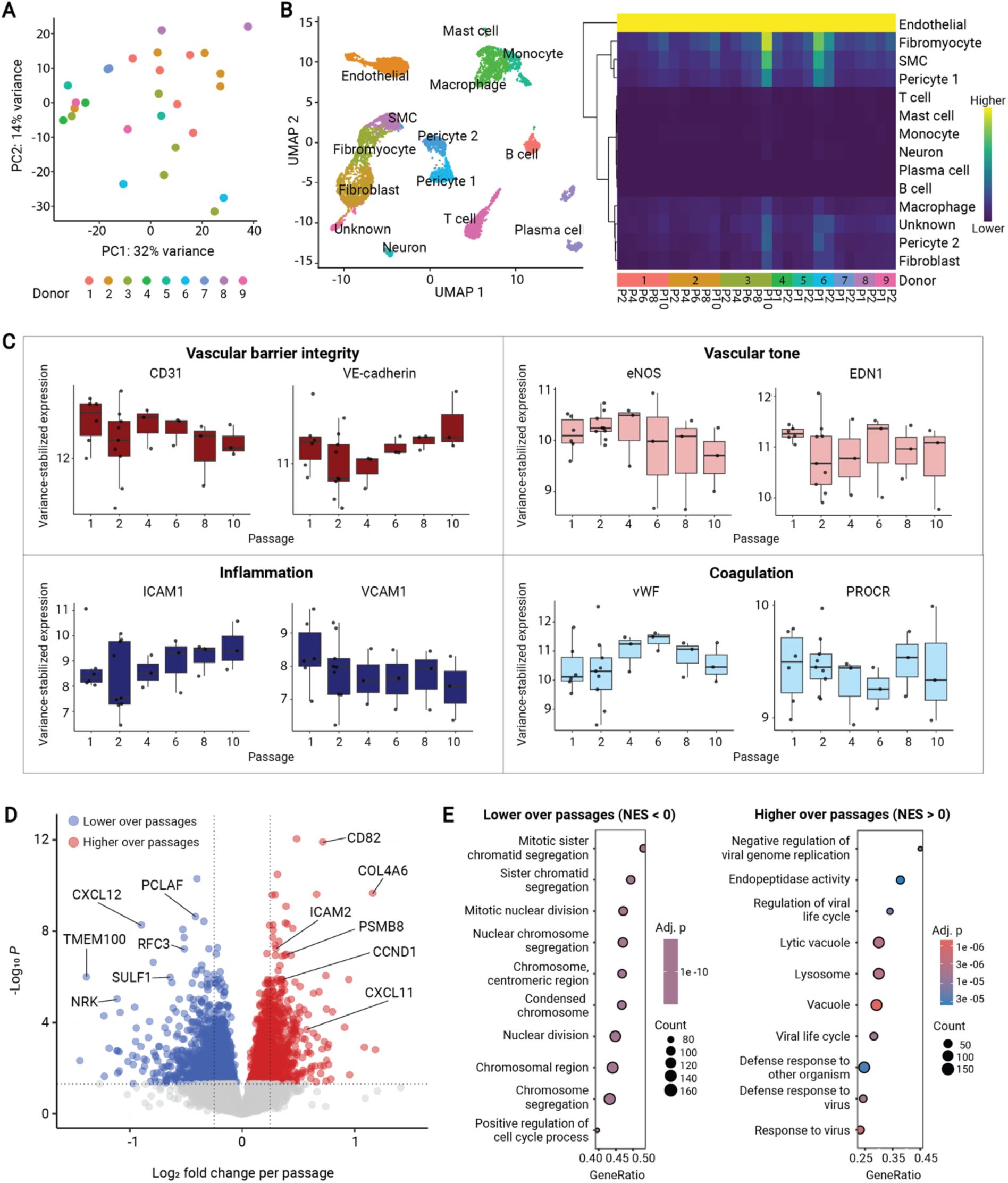
Transcriptional characterization of coronary catheterization-derived endothelial cells (CC-ECs) over passages. A) Principal component analysis of bulk RNA sequencing data from nine donors across passages (P1-P10), showing donor-specific clustering independent of passage. B) UMAP projection onto a human coronary artery single-cell reference atlas confirming localization within the endothelial cluster. C) Stable expression across passages of genes reflecting endothelial function, including barrier integrity (*CD31, VE-cadherin*), vascular tone (*eNOS, EDN1*), inflammation (*ICAM1, VCAM1*), and coagulation (*vWF, PROCR*). D). Differential gene expression analysis modelling passage as continuous variable, identifying genes progressively decreasing (blue) or increasing (red) over time. E) Gene set enrichment analysis shows antiviral/defense-related pathways decrease and cell cycle pathways increase over passages (NES<0: lower over passages; NES>0: higher over passages)

### CC-ECs as cellular model to study endothelial dysfunction

To characterize the suitability of our CC-ECs as an *in vitro* endothelial model, we assessed the functional EC behavior and compared it to HUVECs. First, we performed a scratch assay to evaluate wound healing and cell migration. The wound closure rate between 4 and 16 hours was comparable between CC-ECs (median slope 3.46 (3.43 – 4.23)% per hour) and HUVECS (median slope 4.79 (3.93-4.88) % per hour; p = 1.00) (**Figure 3A**). Second, we assessed angiogenic capacity using a tube formation assay. CC-ECs formed capillary-like networks comparable in morphology to HUVECs. Quantitative analysis revealed no statistically significant differences between CC-ECs and HUVECs in tube length (886 (853 – 896) vs. 874 (867-1089) per mm^2^, p = 1.00), number of junctions (5 (5 – 5) vs. 6 (5 – 9) per mm^2^, p = 0.40) and number of meshes (2 (2 – 2) vs. 2 (2 – 4) per mm^2^, p = 0.70) (**Figure 3B**).

**Figure 3.**
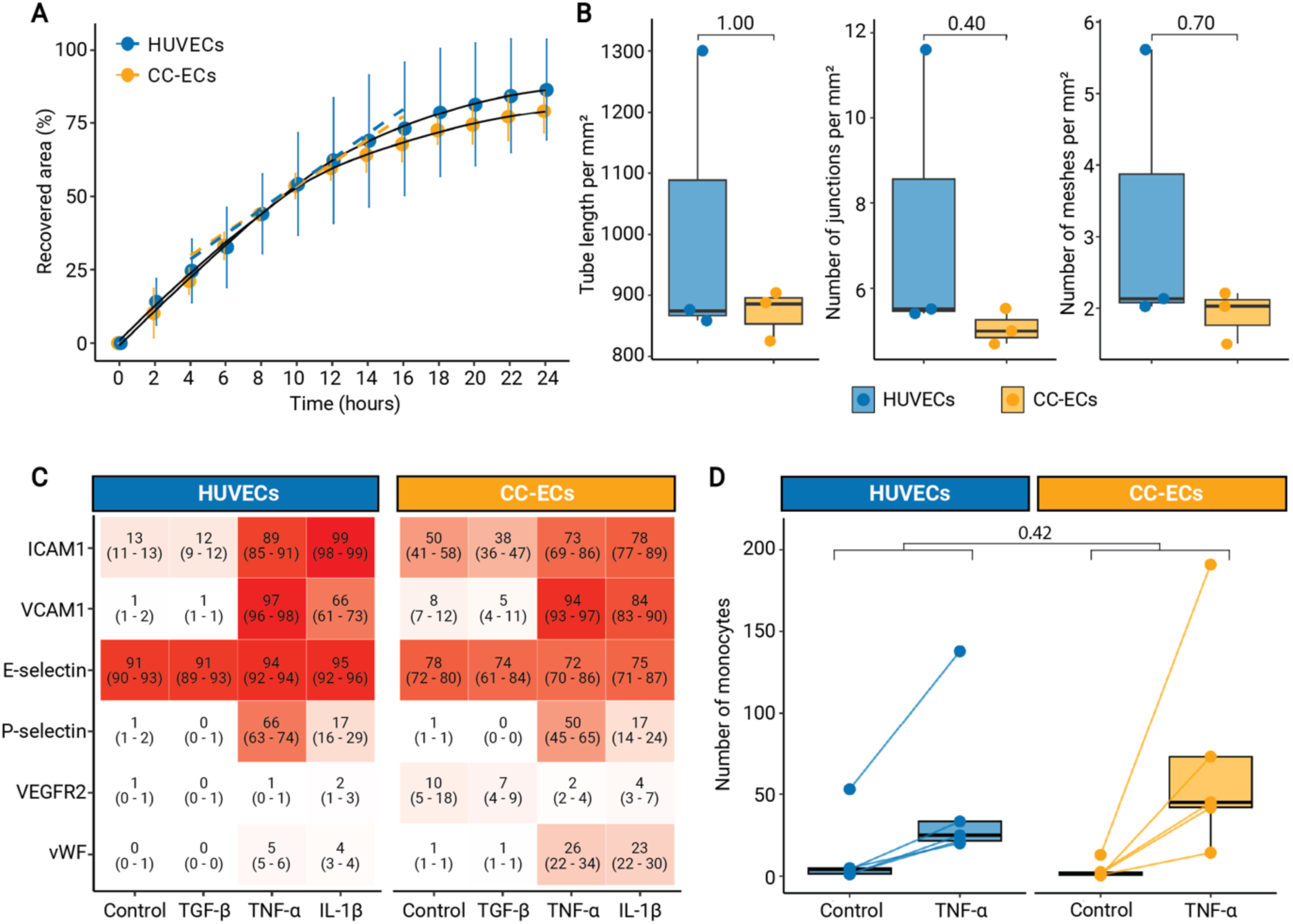
Functional characterization of coronary catheterization-derived endothelial cells (CC-ECs). A) Scratch assay demonstrating comparable wound closure rates between CC-ECs (n=3) and HUVECs (n=3) between 4-16 hours, with linear dashed regression lines indicating the rate of closure. B) Tube formation assay showing similar angiogenic capacity, quantified by tube length, number of junctions and number of meshes per mm^2^ in CC- ECs (n=3) and HUVECs (n=3). C) Heatmap of flow cytometric analysis showing the percentage of marker-positive cells under control conditions and after 24-hour stimulation with TGF-β, TNF-α or IL-1β. Expression of ICAM1, VCAM1, E-selectin, P-selectin, VEGFR2 and vWF was comparable between CC-ECs (n=3) and HUVECs (n=3). Colors and values represent median (IQR). D) Monocyte adhesion under flow conditions assessed using the ibidi system under control and TNF-α-stimulated conditions, demonstrating comparable immune cell recruitment capacity between CC-ECs (n=5) and HUVECs (n=5).

As endothelial dysfunction is closely linked to activation of ECs and subsequent expression of adhesion molecules that regulate immune cell recruitment, we next examined the inflammatory responsiveness of the cells. Following 24-hour stimulation with TGF-β, TNF-α and IL-1β, expression of EC markers (ICAM1, VCAM1, E-selectin, P-selectin, VEGFR2 and vWF) was quantified by flow cytometry. Median expression values are summarized in **Figure 3C**. No statistically significant differences in marker expression were found between CC-ECs and HUVECs under control conditions or following stimulation with TGF-β, TNF-α or IL-1β.

CC-ECs expressed canonical endothelial markers at levels comparable to HUVECs and showed clear upregulation of ICAM1, VCAM1, and P-selectin in response to TNF-α and IL-1β, indicating intact inflammatory responsiveness. Similarly, TGF-β stimulation elicited comparable effects in both EC types. To assess functional consequences of EC activation, we assessed monocyte adhesion using the ibidi flow system under control conditions and after stimulation with 10 ng/ml TNF-α. Monocyte adhesion was comparable between CC-ECs and HUVECs under both control (2 (1 – 2) vs. 4 (1 – 5) monocytes; p=0.42) and TNF-α-stimulated conditions (45 (42 – 73) vs. 25 (22 – 34) monocytes; p=0.42). Furthermore, a similar response to TNF-α was found between CC-ECs and HUVECs (43 (40 – 73) vs. 24 (20 – 29) monocytes; p = 0.42) (**Figure 3D**). Together, these results demonstrate that CC-ECs exhibit inflammatory activation and immune cell-recruitment responses comparable to HUVECs, supporting their functional endothelial behavior.

## Discussion

In this study, we present a refined approach to isolate and culture ECs from catheterization material obtained during routine diagnostic procedures in patients with ANOCA. We demonstrate a successful isolation in approximately half of the included patients, indicating a high degree of reproducibility of the isolation and culturing protocol. Importantly, isolation success was not associated with recorded clinical or procedural characteristics, suggesting that this approach is broadly applicable and translatable to the wider population of patients undergoing coronary function testing. Once established, CC-ECs displayed proliferation rates comparable to HUVECs, expressed canonical endothelial markers and exhibited typical endothelial functional behavior. Gene expression remained stable up until passage 10. Although earlier passages are generally preferred to minimize culture-induced alterations, this stability allows these ECs to be maintained for several weeks, providing a practical window for the development of multiple functional assays.

Our transcriptomic analyses further support the robustness of CC-ECs as a primary endothelial model, while also revealing the effects of prolonged *in vitro* culture. Bulk RNA sequencing across passages showed donor-specific clustering, indicating retention of donor characteristics over time. Projection onto a well-established single-cell reference atlas confirmed stable endothelial identity, with CC-ECs consistently mapping to the EC compartment. At higher passages, however, an increased transcriptional proximity to fibromyocytes and smooth muscle cells was observed, suggesting subtle phenotypic shifts with prolonged culture. Furthermore, several genes that decreased over passages, including *CXCL12*, *TMEM100* and *SULF1*, have previously been linked to endothelial homeostasis, whereas genes that increased with passaging, such as *CD82*, *COL4A6* and *CXCL11*, were associated with extracellular matrix organization and stress response. Although canonical endothelial programs remained largely preserved, these changes may reflect early phenotypic drift, including features compatible with endothelial-to-mesenchymal transition, a process known to be promoted by *in vitro* stress and replicative aging. Importantly, these findings suggest that the use of earlier passages may be preferable for functional studies, particularly when assessing disease-relevant endothelial phenotypes, as they are less likely to be confounded by culture-induced transcriptomic changes.

Functional assays showed that CC-ECs display endothelial behavior comparable to HUVECs, also supporting their use as an *in vitro* endothelial model. Compared with HUVECs, however, CC-ECs exhibited a greater degree of inter-donor variability, most notably in our monocyte adhesion assay. While technical factors cannot be fully excluded, this increased spread may reflect underlying patient-specific differences in endothelial activation. Given the heterogeneous clinical presentation of ANOCA, this variability is expected and highlights the potential relevance of disease-derived endothelial models for capturing inter-individual differences that may not be adequately represented by standard EC lines.

Having established feasibility, this model offers opportunities to investigate disease mechanisms underlying endothelial dysfunction in ANOCA. Patient-derived ECs may enable the study of distinct ANOCA endotypes by allowing assessment of endothelial pathways involved in vascular tone regulation, including nitric oxide signaling, endothelin-mediated vasoconstriction and responses to vasodilatory or vasoconstrictive stimuli. Furthermore, the model may allow testing endothelial interaction with immune cells, which are increasingly recognized as contributors to vascular inflammation and endothelial dysfunction. In addition, this approach could facilitate exploratory studies on endothelial responses to pharmacological interventions, providing insight into inter-individual variability in treatment effects.

At the same time, several limitations should be considered when interpreting the findings of this study. First, we did not systematically record technical factors that may influence EC yield, such as procedure duration or the extent of material manipulation during sampling. Second, cultured ECs are studied outside their native vascular environment and therefore lack interactions with surrounding cell types, particularly vascular smooth muscle cells, which play a critical role in regulating coronary vasomotor function. Moreover, standard static culture conditions do not recapitulate physiological shear stress, nor the complex biochemical milieu of circulating blood. These factors may influence endothelial phenotype and function and should be considered when interpreting *in vitro* findings. Finally, the relatively low number of characterized CC-ECs precluded assessment of potential sex-specific differences in endothelial function. Consequently, the generalizability of our findings to male versus female patients remains uncertain and future studies with larger sample size are needed to address this.

Access to patient-derived endothelium remains a major limitation in vascular research. Previous studies have demonstrated that ECs can be isolated from intravascular devices, including pulmonary ECs obtained from Swan–Ganz catheters, with some reports also showing successful *in vitro* culture^21,22^. In the coronary circulation, a flow cytometry–based approach has enabled isolation and phenotypic characterization of human coronary ECs from guidewires, but without demonstrating their expansion in culture^23^. Building on our prior work^11^, we now show for the first time that ECs isolated directly from routine coronary catheterization material, without balloon-based endothelial contact, can be maintained and expanded *in vitro*. This methodological advance enables functional and transcriptomic analyses of disease-relevant ECs and may be applicable to vascular disease fields with limited access to patient-derived endothelium, including neurovascular disorders (e.g., subarachnoid aneurysms), pulmonary vascular diseases (e.g., pulmonary arterial hypertension) and coronary pathologies (e.g., spontaneous coronary artery dissection).

## Conclusions

In conclusion, this study demonstrates the feasibility of isolating and culturing ECs from coronary catheterization material during routine coronary function testing in patients with ANOCA. The patient-derived cells retain endothelial identity, exhibit donor-specific transcriptional profiles and show functional behavior comparable to established endothelial models, while preserving inter-individual heterogeneity. By enabling access to patient-derived ECs, this approach offers a valuable platform for future mechanistic studies of endothelial dysfunction in ANOCA, with potential applicability across a broad range of vascular diseases.

## Data Availability

Data supporting the findings of this study are available from the corresponding author upon reasonable request.

## Supplemental Material

Supplemental Table S1-S4

Supplemental Figure S1

Data Set

## Non-standard Abbreviations and Acronyms

ANOCA: angina with nonobstructive coronary arteries
CC-EC: coronary catheterization-derived endothelial cell
EC: endothelial cell
HCAEC: human coronary artery endothelial cell
HCMEC: human cardiac microvascular endothelial cell
HDMVEC: human dermal microvascular endothelial cell
HUVEC: human umbilical vein endothelial cell
IL-1β: interleukin-1 beta
MSC: mesenchymal stem cell
TGF-β: transforming growth factor-beta
TNF-α: tumor necrosis factor-alpha
vWF: von Willebrand factor

